# Targeting Optimal Grasp-Related Cortical Areas for Intracortical Brain-Machine Interfaces

**DOI:** 10.1101/2025.10.10.25337598

**Authors:** Tyler R. Johnson, Crispin Foli, Emily C. Conlan, Katherine A. Koenig, Mark J. Lowe, William D. Memberg, Robert F. Kirsch, Eric Z. Herring, Stanley F. Bazarek, Emily L. Graczyk, Dawn M. Taylor, A. Bolu Ajiboye, Jennifer A. Sweet

## Abstract

This study aimed to optimize intracortical microelectrode array implantation sites for grasp-related motor decoding by integrating anatomical, functional, and vascular imaging with preoperative 3D modeling.

A participant with C5 tetraplegia underwent anatomical MRI, diffusion-weighted imaging, and task-based fMRI to identify grasp-related cortical regions while avoiding vasculature and speech-critical areas. Quicktome software was used to refine target selection by integrating structural connectivity and functional activation data. A 3D-printed skull and cortical model enabled preoperative planning, including craniotomy and electrode positioning simulations. Electrode placement was validated postoperatively using neural data collected from the implanted arrays during attempted movements of the arm and hand.

Functional imaging identified distinct grasp-related activation in anterior intraparietal area (AIP), ventral premotor cortex (PMv), and inferior frontal gyrus (IFG). AIP was selected based on its strong connectivity with motor cortex and distinct functional activation. Subregions 6v and 6r of PMv, which exhibited robust grasp-related activity and were surgically accessible, were chosen over the posterior IFG region, which extended into a sulcus making implantation difficult. Postoperatively, the arrays enabled high-fidelity decoding of arm/hand movements, achieving a combined classification accuracy of 96%.

This study presents a multi-modal approach for optimizing intracortical electrode placement by combining MRI-based anatomical mapping, fMRI-guided functional localization, connectivity information, and 3D surgical modeling. These findings demonstrate an effective method for identifying surgically feasible grasp network implant locations in a paralyzed individual. This is an essential step for brain-machine interface systems that use grasp-related brain activity to command devices, such as neuromuscular stimulation systems for restoring upper limb function in individuals with spinal cord injury.

## Introduction

Restoring arm and hand function is the highest priority for individuals with cervical spinal cord injury (SCI), as it directly impacts independence and quality of life.^1–4^ Functional electrical stimulation (FES) has been successfully utilized to reanimate paralyzed muscles, enabling individuals with quadriplegia to generate reaching and grasping movements.^5–9^ However, the effectiveness of FES remains constrained by the limited control options available, particularly for individuals with high cervical injuries who have more limited options for commanding their FES systems.

One approach to bypassing the damaged nervous system is the use of intracortical brain-machine interfaces (BMIs), which decode movement intentions directly from neural activity in the brain.^10–14^ Previous studies have demonstrated the feasibility of BMI-controlled FES to restore movement, using neural signals predominantly from the primary motor cortex to command functional reach and grasp.^5,15,16^ To further advance this methodology, it is necessary to expand beyond the primary motor cortex and implant electrode arrays into additional grasp-related regions of the brain to decode complementary information.^17–19^ This offers the potential to enhance the functionality and adaptability of FES-driven grasp, ultimately improving functional outcomes for individuals with SCI.

Visuomotor control of reaching and grasping is mediated by a distributed network of cortical regions, including the primary motor cortex (M1), dorsal and ventral premotor areas (PMd, PMv), anterior intraparietal area (AIP), superior parietal lobule (SPL), supramarginal gyrus (SMG), and inferior frontal gyrus (IFG).^20,21^ The primary motor cortex (M1) is a key structure for executing hand and arm movements, encoding movement kinematics and dynamics.^22,23^ Components of SPL are specialized for encoding limb movement trajectories and reach planning,^24,25^ while AIP integrates visual information about object shape to facilitate hand pre-shaping during grasping.^26,27^ Premotor areas, particularly PMd and PMv, play a crucial role in planning and coordinating grasp/reach-related motor actions,^28,29^ while SMG supports grasp selection and orientation.^30,31^ Additionally, the IFG contributes to the refinement of grasping and object manipulation, integrating visual and somatosensory input to facilitate precision grip control.^21,32^ These regions collectively support the transformation of visual input into motor commands, enabling dexterous object interaction.

Intracortical microelectrode arrays have been primarily implanted in the primary motor cortex (M1) for movement intention decoding in human intracortical brain-machine interface systems.^11,15,16,33^ However, a limited number of studies have explored electrode implantation in regions specifically associated with reach and grasp control, including the ventral and dorsal premotor cortex (PMv/PMd),^34,35^ the anterior intraparietal area (AIP),^36–40^ the supramarginal gyrus (SMG),^35,36^ the superior parietal lobule (SPL),^18,38,39,40^ and the inferior frontal gyrus (IFG).^34,37^ Due to the anatomical proximity between grasp-related and speech-related brain regions, certain electrode arrays implanted in these areas have been leveraged for both grasp decoding and speech motor decoding.^34,41^

### Challenges in Electrode Implantation

Targeting intracortical electrode arrays for grasp-related motor decoding in individuals with spinal cord injury presents several challenges. First, the two currently FDA-approved intracortical systems, microelectrode arrays from Blackrock (Blackrock Neurotech, Salt Lake City, UT) and flexible, individually inserted threads from Neuralink, share a significant limitation. Despite their different designs, both are primarily implanted into the exposed surfaces of the cortex (gyri). This constraint restricts implantation to exposed cortical surfaces, thereby excluding sulcal regions such as the primary motor and sensory cortices within the central sulcus, the anterior intraparietal area (AIP) within the postcentral and intraparietal sulci, and portions of the ventral and dorsal premotor cortex (PMv/PMd) located within the precentral sulcus.

Additionally, the complex and highly individualized anatomy of cortical sulci and gyri further complicates consistent targeting across participants. While broad structural landmarks are conserved across individuals, their specific morphologies vary substantially, akin to a fingerprint.^42,43^ This inter-individual variability is exacerbated in spinal cord injury, as cortical atrophy and neuroplastic reorganization following injury can further distort expected structural-functional relationships, making it more difficult to reliably identify functionally equivalent implant sites across individuals with anatomical landmarks alone.^44,45^

Targeting non-primary motor regions, particularly the AIP area, presents a significant challenge due to substantial inter-individual variability in functional localization. Although sulcal landmarks offer a general anatomical guide, the precise location of grasp-related activity can vary markedly from person to person, increasing the risk of misplacing electrodes into neighboring, functionally distinct areas when relying on anatomy alone. While primary sensory cortex is often a target for sensory BMIs and can be readily mapped intraoperatively, non-primary sensory areas, such as AIP and IFG, do not elicit clear sensory responses to intraoperative electrical stimulation and are therefore challenging to functionally define during a surgical procedure. Pioneering studies have demonstrated the feasibility of implanting arrays in high-level association cortices, specifically posterior parietal regions, to decode cognitive aspects of movement goals.^18,38–40^ Resulting implant locations from these studies have also brought to light the considerable variability in the functional regions across individuals (Supplementary Figure 1). Our own prior work reinforces this understanding: an array implanted in AIP based on anatomical landmarks exhibited unexpected sensitivity to auditory stimuli, emphasizing the complex functional topography of the parietal cortex (unpublished data).^37^ Together, these findings highlight the critical importance of complementing anatomical targeting with individualized functional mapping to ensure arrays are optimally placed to capture grasp-related neural activity.

As mentioned, many grasp-related cortical regions are located in close proximity to speech-related areas, which are especially critical for individuals with severe motor impairment given that they enable communication. Thus, surgical planning must carefully balance access to motor-relevant regions with the necessity of preserving speech function. Even after identifying the most suitable cortical surface targets, surgical constraints impose additional limitations. Neurosurgeons must also navigate around major vasculature, select implant locations compatible with craniotomy constraints, and ensure sufficient space for connecting electrode leads, skull-mounted pedestals, and reference/ground wires, all of which further constrain possible implantation sites.

### Approaches For Identifying Cortical Targets

To address these challenges, several approaches have been employed to identify functionally and anatomically optimal cortical targets for reaching and grasping-related motor decoding while ensuring safety and accuracy. General cortical anatomy provides an initial framework for target selection, leveraging major landmarks such as the central, postcentral, lateral, and intraparietal sulci to delineate regions of interest. This anatomical approach can be further refined by registering an individual’s brain morphology with population-based cortical geometry databases (e.g., Human Connectome Project [HCP]), allowing for a statistically guided estimation of functional areas. However, grasp-related cortical regions remain less well defined anatomically, necessitating additional functional localization techniques.^35,38^

Functional mapping can further enhance target identification by directly assessing cortical activity during relevant tasks. Functional magnetic resonance imaging (fMRI) has been used to localize motor areas,^11,15,20^ sensory areas,^19,46^ and even reaching/grasping-related regions for microelectrode targeting.^38,39^ Given the considerable inter-individual variability in brain anatomy, functional mapping provides individualized localization of task-specific areas, reducing reliance on anatomical or population-based predictions alone. This is particularly valuable for individuals with tetraplegia, where the inability to physically perform motor tasks makes determining cortical activation for those tasks more challenging.

In addition, intraoperative electrophysiological mapping, including direct cortical stimulation (DCS), provides real-time validation of sensory and motor targets, particularly in somatosensory areas where elicited percepts can be confirmed by the patient using awake surgical techniques.^37,47^ Though not without risk, these techniques are particularly valuable in cases where cortical plasticity or injury-induced reorganization may have altered the expected functional representations.

### Current Study Approach

In the present study, we utilized multiple strategies to identify functionally and anatomically optimal cortical targets for implanting electrodes to decode reaching/grasping while ensuring surgical feasibility and safety. We employed anatomical classification based on Quicktome registration (Omniscient Neurotechnology; Sydney, Australia), which integrates data from the Human Connectome Project (HCP)^48^ and tractography analyses to approximate cortical regions based on structural and functional connectivity. To further refine target selection, we implemented an ‘imagined movement’ fMRI protocol to identify regions selectively activated during reaching and grasping. Additionally, we conducted an auditory fMRI protocol to localize speech and auditory processing areas, ensuring exclusion of these regions from potential electrode implantation sites to preserve communication. Beyond functional and anatomical mapping, we incorporated contrast-enhanced MRI scans to better define safe corridors for cortical access between the overlying vasculature. To optimize surgical planning, we generated a 3D model of the participant’s skull and cortex, enabling preoperative simulations of craniotomy placement and array positioning. This enhanced ability to integrate the required surgical steps improved procedural accuracy and reduced surgical time and intraoperative risks. Finally, we evaluated the efficacy of the placement of the electrodes by observing the neural responses as recorded by the electrodes during attempted arm and hand movements.

## Materials and methods

### Participant

Participant RP2 (participant ID assigned only within this research), a male with C5 AIS B tetraplegia, was enrolled into the study (ClinicalTrials.gov ID: NCT03898804). An Investigational Device Exemption was obtained from the U.S. FDA for the study. The electrode implantation and testing protocol was reviewed and approved by the University Hospitals Institutional Review Board. Informed consent was obtained for study enrollment and prior to all procedures. Additional fMRIs were also collected from two additional able-bodied individuals as part of a separate imaging study protocol approved by the Cleveland Clinic Institutional Review Board after a separate informed consent process.

### Anatomical Mapping

Three months prior to surgery, RP2 underwent standard MRI scans including T1-weighted, T2-weighted, and FLAIR sequences, along with diffusion-weighted imaging and high-resolution volumetric imaging with gadolinium-enhanced T1-weighted scans. A high-resolution CT was also obtained to assist with surgical planning. Using these scans, major landmarks were located, and preliminary cortical targets were identified along with prohibitive vasculature. Skull and cortical models were extracted from these scans as detailed below.

The MRI scans were then imported into Quicktome, where anatomical areas of interest were further identified and mapped using a multi-modal approach. Quicktome provides two complementary methods for identifying and refining brain regions. The first method leverages the Human Connectome Project Multi-Model Parcellation (HCP-MMP1) atlas as a reference for brain anatomy. By aligning the participant’s unique cortical geometry, including gyri and sulci contours, with this atlas, Quicktome applies advanced machine learning algorithms and constrained spherical deconvolution (CSD)-based tractography to generate a highly personalized brain map. This approach not only accounts for individual anatomical variations but also compensates for distortions caused by pathology, such as atrophy or structural reorganization, ensuring highly accurate region identification. We utilized these software features to identify anatomical regions of the participant’s brain for further analysis.

The second method focuses on functional connectivity analysis, utilizing resting-state fMRI data to assess correlations in neural activity across brain regions. By extracting time-series data from predefined parcels and computing correlation matrices, Quicktome identifies co-activation patterns that reveal functional networks and refine region specificity. This dual approach, combining precise structural alignment with the Human Connectome Database and dynamic functional connectivity mapping, provides a comprehensive, individualized understanding of the participant’s brain anatomy, enabling precise localization of areas of interest. This connectivity analysis was used to locate regions of interest along with their relationship with other anatomical regions of interest, helping to ensure that we were able to find targets in the reaching/grasping network of the cortex.

### Task-Based Functional Mapping

An imagined grasp/reach fMRI task was used to localize relevant motor areas. Scanning was conducted on two able-bodied test subjects before scanning the primary participant. These preliminary studies allowed for refinement of task instructions and optimization of analysis procedures. The resulting modifications enhanced data quality and streamlined workflow efficiency. Images from both able-bodied subjects and the primary SCI participant are included in this report.

### fMRI Data Acquisition

Scans were conducted using a Siemens 3T MAGNETOM Prisma Fit scanner equipped with a 32-channel head coil. Structural images were acquired using an MPRAGE sequence (*TR* = 2.3 s, *TE* = 2.98 ms, *flip angle* = 9°, *matrix size* = 256 × 256). Functional images were obtained using an echo-planar BOLD sequence with a multiband factor of 3, *TR* of 1.7 s, *TE* of 30 ms, *flip angle* of 65°, and a *matrix size* of 104 × 104.

The imagined grasp/reach task was based on previously published paradigms,^38,39^ and adapted to enhance the signal-to-noise ratio. Each block began with the presentation of a two second cue depicting one of three grip types: power grip, pinch grip, or reach. This cue directed participants on how to mentally interact with the subsequent stimulus. Following this cue, a blue/green bar appeared for 3 seconds in one of six different spatial locations and participants were instructed to imagine reaching towards the bar or grasping it using the designated grip. A 10-second rest period signaled the end of the trial before the cycle was repeated. Figure 1 shows the stimuli and procedure for each block. For each scan, eight “reach” blocks and eight “grasp” blocks were presented. The pinch and power grip conditions were both considered “grasp” blocks and were analyzed together. The scan session included three eight-minute runs of this task. Participants were instructed to remain completely still throughout the scan to minimize motion artifacts. They were asked to maintain focus on the tasks to ensure optimal activation of the target brain regions.

**Figure 1.**
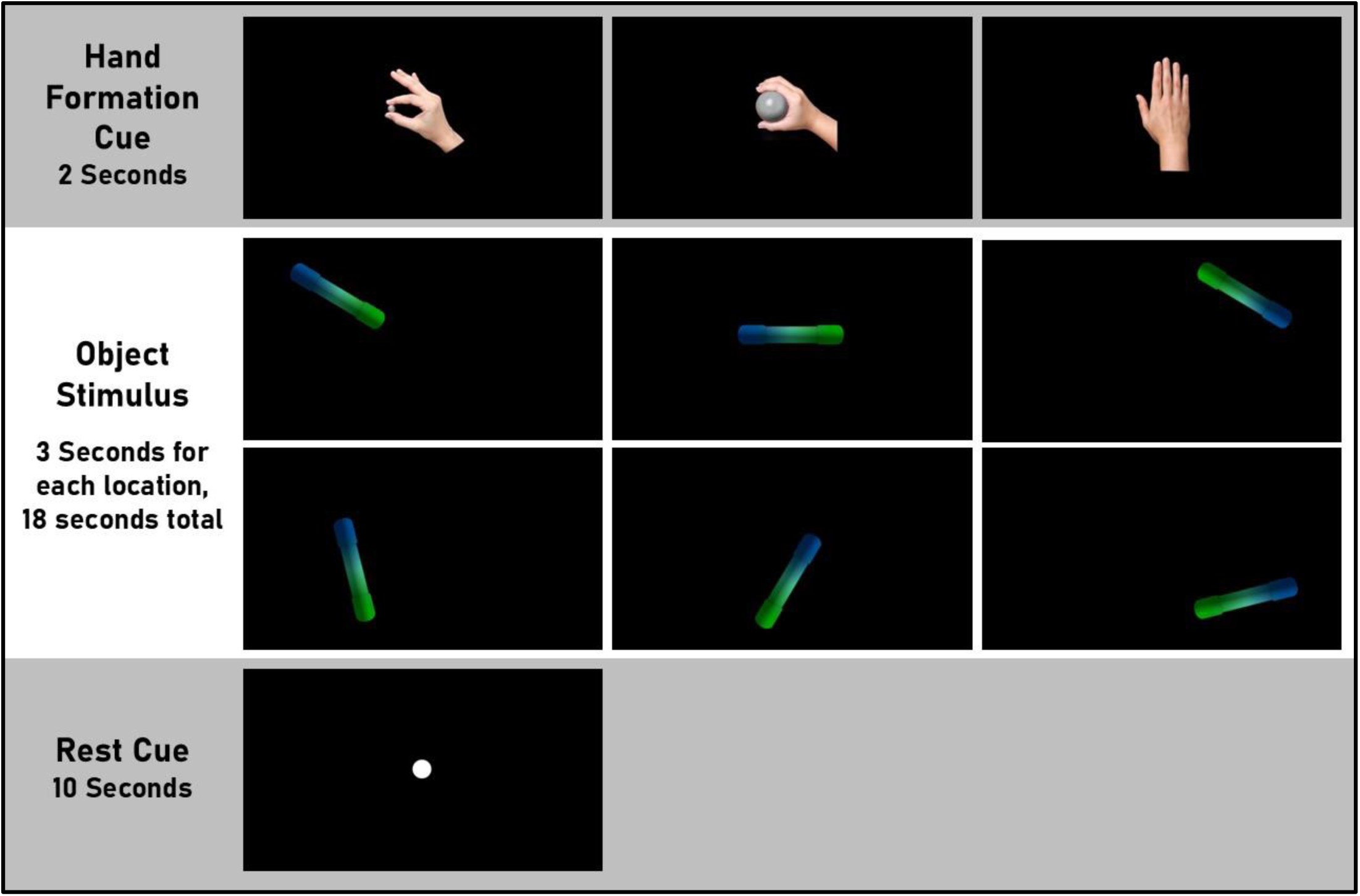
Imagined grasp/reach block illustration. The top row shows the three possible hand cues (pinch grip, power grip, reach). The middle rows show the six positions of the stimulus object. The bottom row shows the rest visual cue.

Auditory activation was measured using a 5-minute block design paradigm during which the participant was instructed to lie still and listen as blocks of reversed speech were alternated with blocks of silence. This auditory paradigm was selected based on prior pilot testing, demonstrating high reproducibility in identifying sound-responsive brain areas across multiple sessions. Participant RP2 was the only individual to undergo both the audio fMRI and the imagined grasp/reach task.

### fMRI Analysis

#### Data Preprocessing

All preprocessing was performed using Analysis of Functional NeuroImages (AFNI),^49^ Surface Mapper (SUMA),^50^ and the FMRIB Software Library within a Linux environment.^51^ The first four volumes of each fMRI time series were removed to account for T1 equilibration effects. Preprocessing included slice timing correction, motion correction, spatial alignment, and smoothing. Motion correction was performed using Slice-Oriented Motion Correction,^52^ which applies slice-wise motion regression to minimize motion-related artifacts. For the imagined grasp/reach movement scans, runs 2 and 3 were aligned to run 1 using AFNI, ensuring spatial consistency across runs. The anatomical T1-weighted image was aligned to run 1 to serve as a reference for cortical surface projection. Spatial smoothing was applied using a 4 mm full-width at half maximum Gaussian kernel to improve signal-to-noise ratio. Similar methods were also applied to the auditory task imaging data.

#### fMRI Analysis and Statistical Modeling

Task-related activation was identified using a general linear model (GLM) implemented in AFNI’s 3dDeconvolve. Task timing was extracted from stimulus logs and structured into separate files corresponding to grasp and reach blocks. Hemodynamic responses were modeled using AFNI’s BLOCK function with a duration of 18 seconds per event (3 seconds x 6 target locations). A contrast of grasp versus reach was computed to assess differences in activation patterns. The resulting statistical maps were thresholded using a voxel-wise threshold of *p*<1.0×10-5 and a *cluster-size* of 100 to account for multiple comparisons.

#### Cortical Surface Projection

The aligned anatomical T1-weighted image was processed using the FreeSurfer recon-all pipeline.^53^ Functional data were aligned to the individual cortical surface reconstruction, enabling visualization of activation patterns across the cortex. Cortical surfaces were visualized using SUMA. This pipeline allowed for the identification of brain regions involved in imagined grasping and reaching movements and auditory processing. The final statistical maps revealed differential activation patterns associated with the task conditions, projected onto cortical surfaces for anatomical interpretation.

### Awake Stimulation Mapping

Regions of cortex were also queried for function with direct cortical stimulation during the implantation procedure. Stimulation was applied with an Ojemann stimulator with current applied at 5 mA. The participant was awoken from anesthesia for approximately 30 minutes to provide verbal responses. The stimulator probe was first applied to regions of the primary somatosensory cortex to identify the target locations for the S1 implants. After each stimulation application, the participant reported which regions of the hand and fingers were perceived. The surgeon marked each probe position eliciting sensation with a paper marker, and after testing was complete, selected the optimal set of positions that elicited sensation over diverse, functional regions of the hand while avoiding blood vessels. Second, cortical stimulation was applied to ensure that the targeted areas were not implicated in speech. Stimulation up to 10 mA was applied while the participant verbally counted to twenty, listed the letters of the alphabet, and moved his tongue. Speech arrest was monitored during counting and alphabet naming tasks. Disruption of tongue movement during stimulation was used to test for speech apraxia. Probe positions that arrested speech or tongue movement were not selected for array placement.

### Surgical Practice using Physical Models

#### Skull Model Fabrication

A high-fidelity 3D-printed skull model was generated from MRI and CT imaging data using 3D Slicer.^54^ The skull was segmented using the Segment Editor module, with additional refinements performed to fill gaps and remove minor artifacts (Fig. 2A,B). To facilitate integration into a surgical mount while maintaining access to the cranial cavity, the model was sectioned in the axial plane, preserving the frontal bone while truncating at the intersection with the zygomatic and nasal structures.

**Figure 2.**
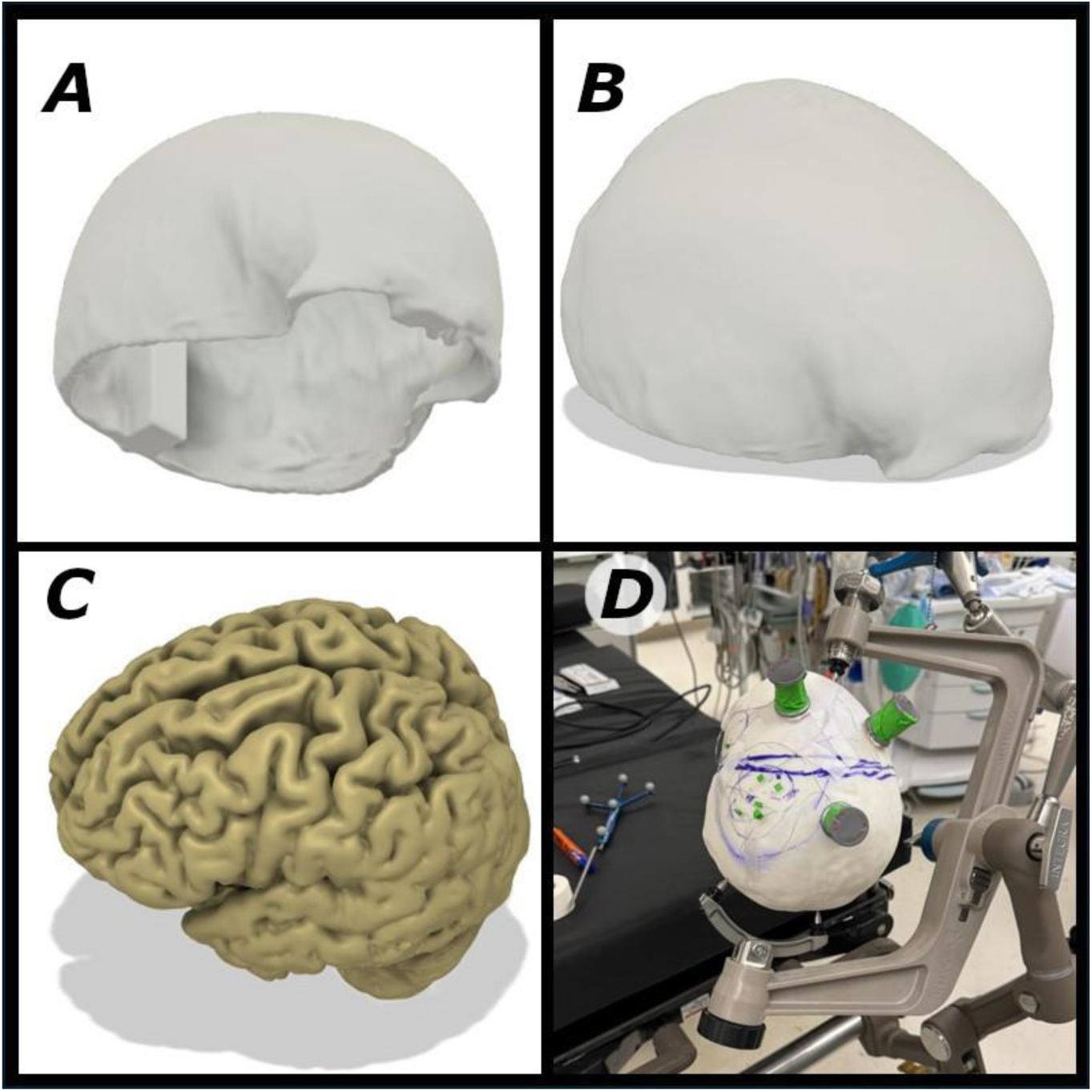
3D models used for surgical planning and practice. **(A)** Underside of skull model showing rectangular protrusion for alignment. **(B)** Skull model side view. **(C)** Cortical model. **(D)** Surgical simulation using skull/cortical models.

The final skull model was exported and printed at true anatomical scale using a Stratasys J750 Digital Anatomy 3D Printer. BoneMatrix™, a photopolymer material engineered to mimic the mechanical properties of human bone, was used for fabrication. The model incorporated a rectangular alignment feature near the occipital region, designed to precisely position the 3D cortical model within the cranial cavity (Fig. 2A).

### 3D Brain Model Fabrication

A 3D brain model was generated following a similar workflow, but rather than directly printing the model brain, a negative mold of the cortical surface was created. Using 3D Slicer, the cortex was segmented from the MRI data (Fig. 2C), and a shell structure was formed around its exterior surface to serve as a mold. Like the skull model, the mold included a rectangular protrusion to ensure proper alignment within the skull model.

The cortical mold was printed using a PolyJet 3D printer with a standard transparent photopolymer resin. After fabrication, the mold was used to cast a cortical model using a curable silicone rubber, selected for its biomechanical properties resembling cortical tissue. This silicone model was subsequently positioned within the printed skull, ensuring anatomical accuracy to allow the neurosurgeon to realistically practice the entire electrode implantation procedure including making the craniotomy, implanting the electrodes, routing the wires, and mounting the percutaneous connectors to the skull.

### Post Implantation Neural Activity Characterization

Neural data collection from the eight implanted Blackrock electrode arrays started four weeks after surgery to investigate the response of the cortical recordings to intended movements. Neural data was based on the “whole-body map” procedures described by Willett.^55^ Neural data were recorded from all 384 channels (M1, S1, AIP, 6V, and 6R) while the participant was presented with a randomized selection of one of 42 experimental conditions covering a broad range of representative movements, along with a single “Do Nothing” cue, per trial. Each experimental day consisted of 10-20 repetitions of each of the 43 total conditions, with one to two repetitions collected per 5–10-minute block of data.

The participant was shown three distinct epochs during each trial: cue, move, and return. During the cue epoch, the participant was instructed via the computer screen about the upcoming condition. The words “Prepare:” and the condition name were presented, along with a picture of the action outlined in red. During the move epoch, the participant was instructed to attempt the movement a single time, regardless of physical ability to complete it. The onset of this epoch was indicated by the word “Go” on the screen, with the condition image shown with a green border. The participant was instructed to isolate each movement as much as possible, leaving the rest of the body still. During the return epoch, the participant was instructed to return to a neutral sitting position, shown on the screen as “Return” with a green box beneath it. This process is illustrated in Figure 8A. Epoch durations were as follows: cue randomized between 2 and 3 seconds; move fixed at 3 seconds; and return fixed at 2 seconds.

#### Neural Signal Acquisition and Preprocessing

Broadband neural activity was recorded at 30 kHz using the NeuroPort system (Blackrock Neurotech, Salt Lake City, UT). Signals were bandpass filtered between 0.25–5 kHz using a zero-phase, fourth-order Butterworth filter. Common average referencing (CAR) was applied on an array-wise basis to mitigate common-mode noise. Spike band power (SBP) features were extracted by computing the mean squared signal amplitude within non-overlapping 10ms bins.^12^ To account for session-wise variability and long-term drifts, SBP features were z-scored on a block-by-block basis. Finally, features were smoothed using a Gaussian kernel with a 50 ms standard deviation.

#### Decoding of Hand/Arm Movements

Nonlinear multi-class support vector machines (SVMs) (using an RBF kernel) were trained to decode the five movement types from the SBP features. To quantify movement-specific encoding, we trained lightweight support vector machine (SVM) classifiers to predict movement type using SBP features from one electrode at a time. Classification was performed on the mean SBP values within a 2-second task window, using electrode subsets optimized per array. Optimal subsets were selected using a custom greedy forward selection algorithm summarized as follows^56^:

I. Initialize set with the electrode that yields the highest individual decoding accuracy.
II. Iteratively add the electrode that, when combined with the current set, minimizes decoding loss (i.e., either yields the greatest improvement–or least degradation– in decoding accuracy)
III. Repeat (from step II) until maximum number of electrodes is reached
IV. Identify the electrode subset that produces the most accurate decoding results for each array

This procedure reduces overfitting by excluding redundant channels and noise contributions, resulting in more compact and informative feature sets. The number of electrodes selected, and the stepwise accuracy improvements are shown in Supplementary Figure 2. Classification performance was evaluated using 10-fold cross-validation, and statistical significance was assessed by comparing decoding accuracy to a chance-level baseline of 1/*C* (where *C* = number of movement conditions).

## Results

### Anatomical Mapping

Following the creation of a three-dimensional (3D) surface model of the participant’s cortex, major anatomical landmarks, including the central sulcus, postcentral sulcus, precentral sulcus, intraparietal sulcus, lateral sulcus, and inferior frontal sulcus, were identified and used to delineate key cortical regions. These landmarks facilitated the localization of the precentral gyrus, postcentral gyrus, inferior frontal gyrus, and superior parietal lobule.

Next, the anatomical results from the Quicktome analysis were mapped onto the cortical surface, allowing for the visualization of functional connectivity between candidate implantation areas (Fig. 3A,B). Additionally, Quicktome-generated parcellation data, which shows anatomical regions based on standard anatomical data, and tractography models were incorporated to support precise localization and connectivity assessments (Fig. 2C,D). The connectivity map in Figure 2E quantifies the scale of the functional networks identified by Quicktome (as demonstrated in Fig. 2A,B). This connectivity analysis helped inform implant targeting by highlighting regions with strong connectivity to motor-related areas likely involved in grasping and reaching.

**Figure 3.**
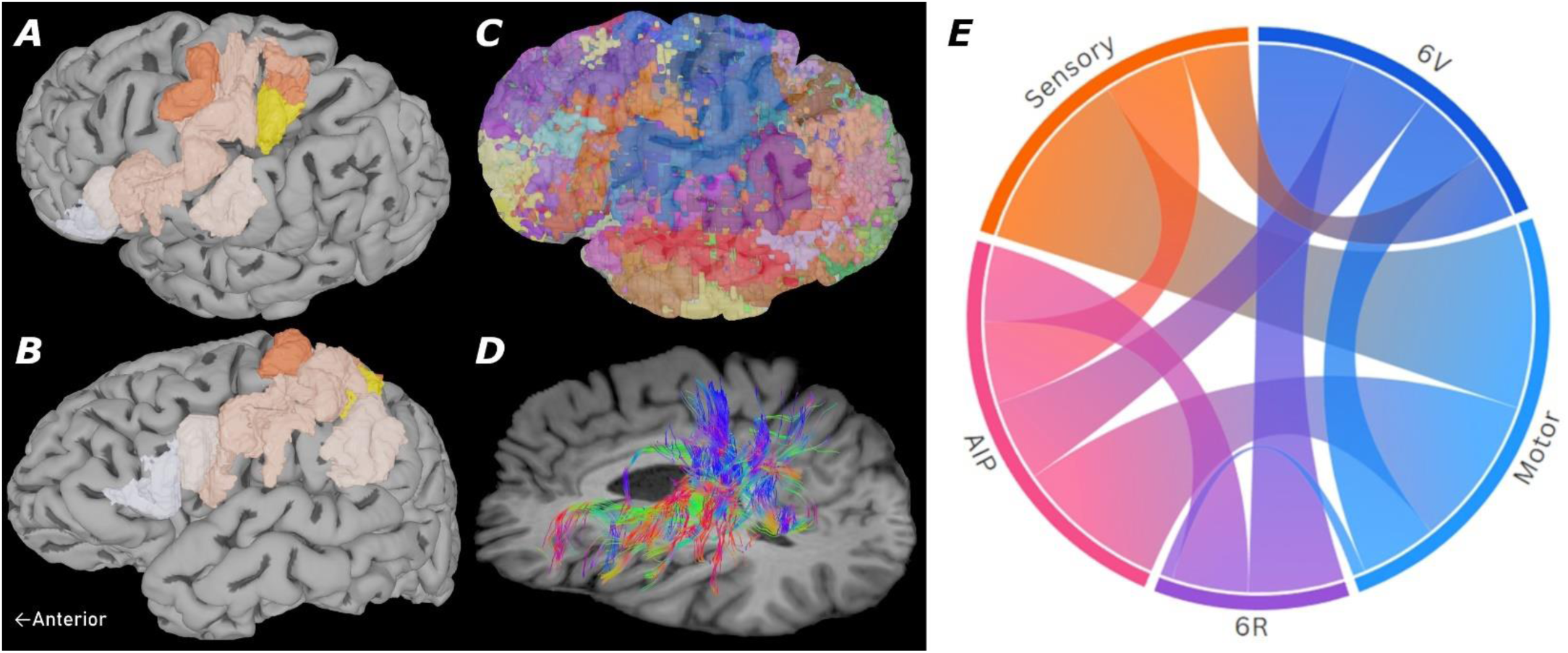
Imaging-based anatomical and connectivity models. **(A and B)** example Quicktome functional connectivity results (white and orange) showing connectivity with AIP (yellow) as indicated by the color intensity (darker orange = more connectivity). **(C)** Quicktome generated parcellation of database-predicted anatomical regions. **(D)** Quicktome generated tractography between anatomical regions of interest. **(E)** Statistical connectivity based on MRI resting-state activity correlations across relevant parcellations from Quicktome where width of connected bands represent correlation across anatomical regions.

### Task-Based Functional Mapping

Results from the imagined grasp/reach task are presented in Figure 4 for both able-bodied test subjects and the primary participant. The first column displays activation during the grasp condition (pinch grip or power grip), the second column shows activation during the reach condition, and the third column shows the difference between the two, obtained by subtracting reach from grasp. In the first and second columns, warm colors indicate positive activation, and cool colors indicate negative activation. In the third column, warm colors indicate stronger activation during the grasp condition and cool colors indicate stronger activation during the reach condition. Images in the reach and grasp columns have been thresholded at *t* = 4.5 with a minimum cluster size of 100 voxels to enhance statistical reliability. Images in the grasp>reach column have been thresholded at *t* = 3.5. Overall, grasp and reach activations were largely similar, yet the contrast showed significant differences, with test subject 1 and the patient showing consistently stronger grasp activation in subregions of the primary sensory and primary motor cortices.

**Figure 4.**
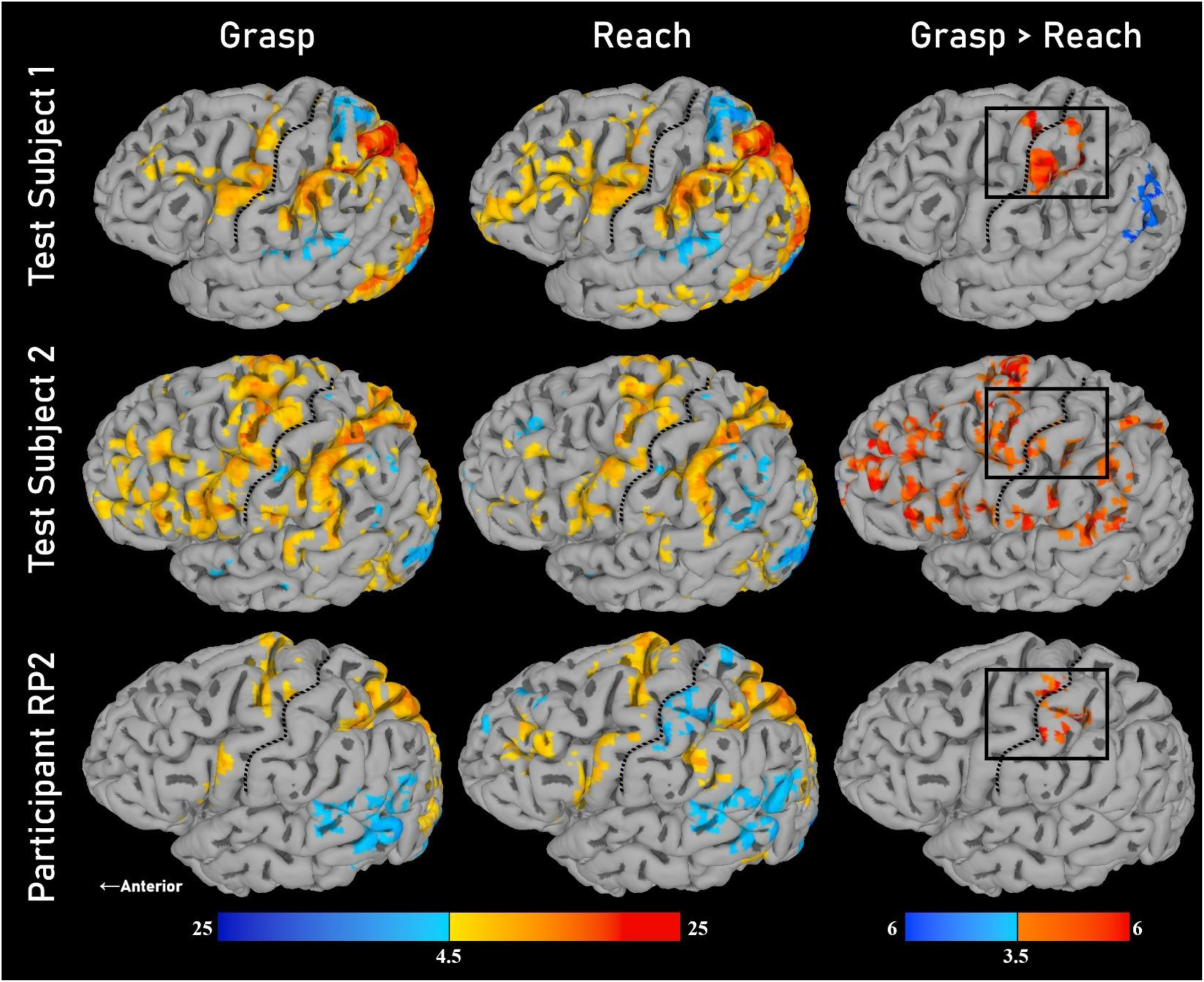
fMRI activation results. Results for able-bodied subjects (top and middle rows), and the SCI cortical implant participant (bottom row). The first column displays brain activation during grasping tasks (pinch grip or power grip), the second column shows activation during reaching, and the third column represents the difference between grasping and reaching activation, obtained by subtracting reaching-related activation from grasping-related activation. In the first two columns (thresholded at *t*=4.5), yellow, orange, and red regions indicate increased blood flow during the task condition compared to rest, with red representing the highest level of activity, whereas blue regions denote decreased blood flow. For the ‘Grasp>Reach’ column (thresholded at *t*=3.5), the color scale represents the difference between grasp activation and reach activation with yellow/red colors indicating stronger grasp activation and blue colors indicating stronger reach activation. The boxes in the third column highlight areas of higher grasp activation for Test Subject 1 and RP2. All images have a *cluster size* of 100 voxels to enhance statistical reliability. Central sulcus is shown as a black dotted line in all images.

### Audio fMRI

The comparison between grasping and audio stimulation highlighted mutually exclusive activation areas providing potential target locations that would record grasp/reach related activity without confounds from speech activity.

### Implant Locations

Target electrode implant locations were selected by integrating cortical anatomy, Quicktome imaging, vascular constraints, awake stimulation, and functional activation during both audio and grasping tasks. In addition to standard primary motor and sensory targets, our initial set of candidate regions within the grasping network included the anterior intraparietal area (AIP), ventral premotor cortex (PMv), and inferior frontal gyrus (IFG).

AIP was first identified anatomically at the medial-posterior intersection of the intraparietal and postcentral sulci (Fig. 6). Quicktome’s functional connectivity analysis revealed a strong connection between this area and the hand knob region of the primary motor cortex, further supporting this target (Fig. 3A,B,E). fMRI results demonstrated grasp-related activation within a subregion of AIP, which was distinct from an adjacent subregion activated by the audio task (Fig. 5). This distinction was critical for precise targeting, as without fMRI guidance, we might have incorrectly selected the more anterior, audio-related portion of AIP. Given the presence of large vasculature near this site, the grasp-related AIP subregion immediately posterior to the vessel, which also matched with the grasp-related activation from the fMRI, was ultimately selected for implantation (Fig. 6D).

**Figure 5.**
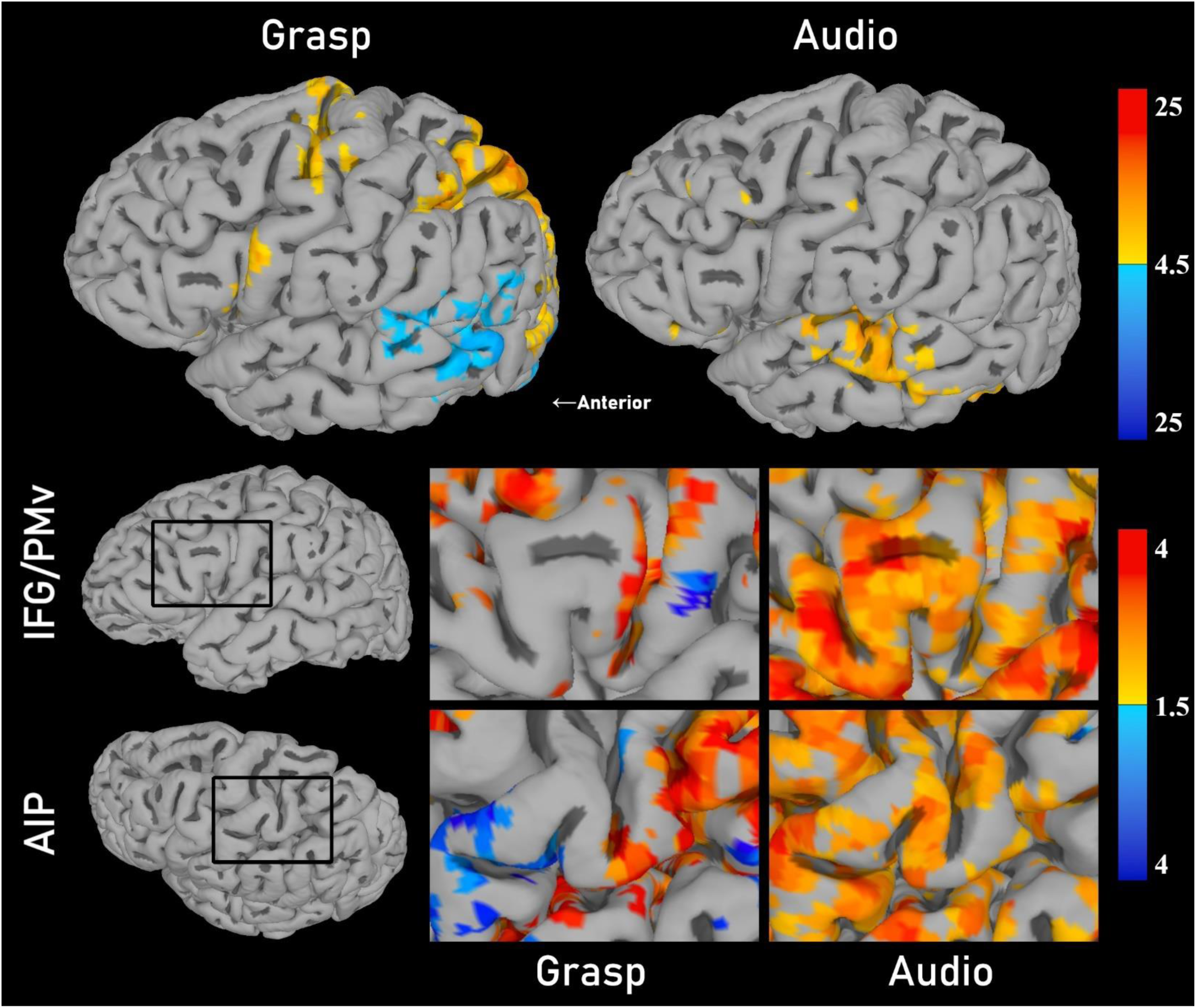
Audio vs. grasp task-based functional mapping results. Top) Participant RP2’s cortical fMRI activation for the grasp condition and audio condition (thresholded at *t*=4.5). Bottom) Enlarged/rotated views highlighting IFG/PMv (top row) and AIP regions (bottom row) for both the grasp condition (left column) and audio condition (right column). These enlarged images have been thresholded to *t*=1.5 to compare areas more associated with grasp vs. audio conditions despite not meeting the *t*=4.5 threshold.

**Figure 6.**
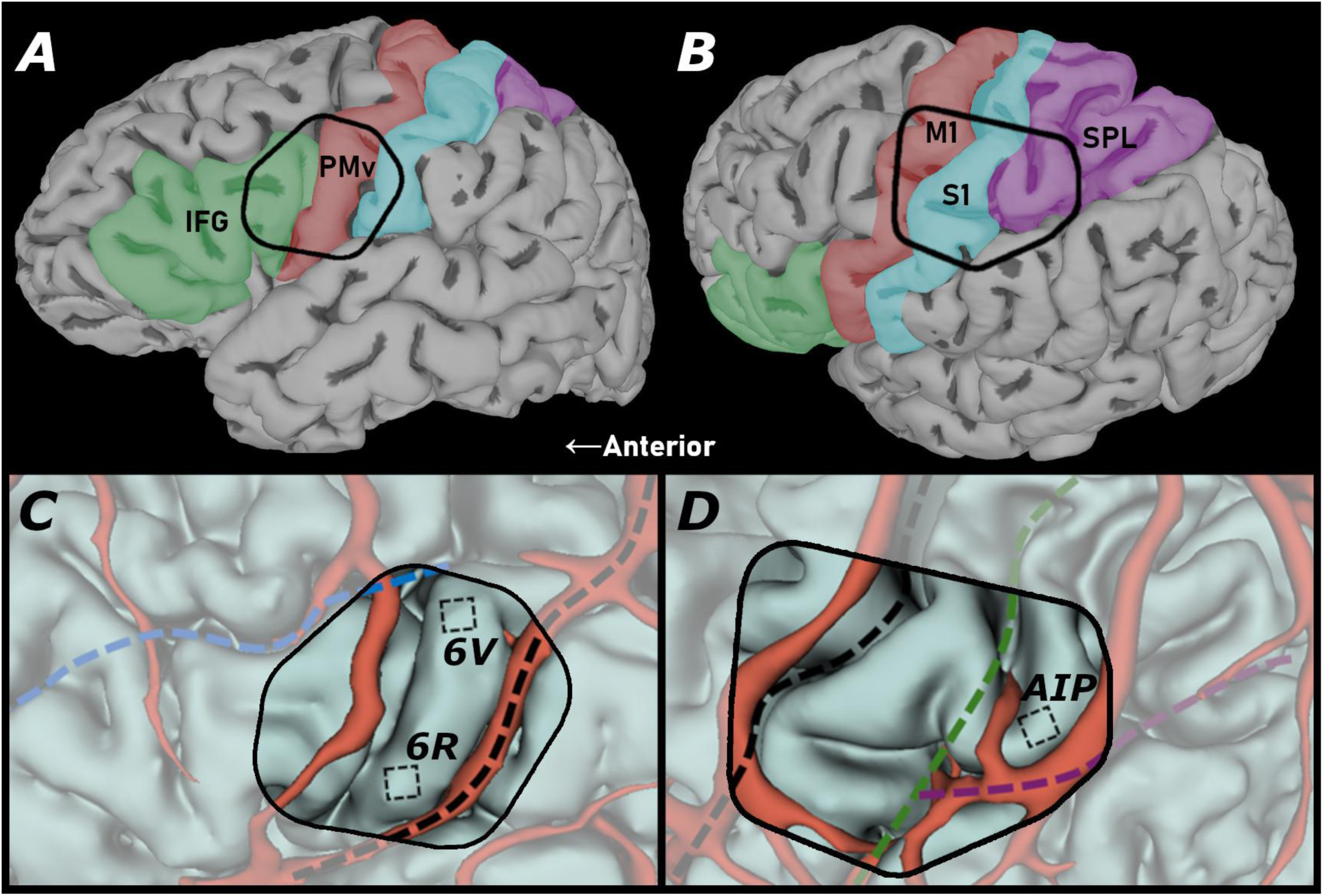
Implant location planning based on anatomy and vasculature. (A and. **B)** delineations of cortical regions based on anatomical scans centered at regions of interest (IFG = green, PMv/PMd = red, M1 = blue, SPL = purple). **(C and D)** cortical models with proposed implant sites and potentially interfering vasculature (central sulcus = black, postcentral sulcus = green, intraparietal sulcus = purple, inferior frontal sulcus = blue)

A similar pattern was observed in the IFG, where Brodmann area 44 exhibited grasp-related activation specifically in its posterior subregion, while the remainder of IFG was predominantly active during the audio task, as expected (Fig. 5). However, this posterior subregion of area 44 extended into the sulcus and the surface component was largely occluded by vasculature, making it a challenging implantation target (Fig. 6C). Alternatively, PMv demonstrated grasp-related activation, superior accessibility for implantation, and an absence of interfering vasculature (Fig. 5,6C). Although PMv in general exhibited weaker functional connectivity with the motor cortex hand region (Fig. 3E), prior research has established its role in grasp-related processing.^34,35^ Thus, we selected both the anterior-dorsal region of PMv (6v) and the anterior-ventral region of PMv (6r) as the final implantation site, as depicted in Figure 6C.

Additional intracortical microelectrode arrays were implanted in primary motor and primary somatosensory using surgical procedures as previously described.^5,37^ Primary sensory and motor targets were identified on either side of the central sulcus with motor locations more medial. Sensory arrays were rectangular instead of square to give more space between individual channels increasing the range of tactile locations likely to be activated with stimulation through these sensory arrays. Since the leads stemmed from the shorter side of these rectangular arrays, it was decided to orient these arrays perpendicular to the central sulcus to allow for the arrays to be placed close together without leads from one array overlapping the neighbouring array which could have prevented the arrays from being fully inserted and seated against the cortex.

### Confirmation of Array Placement in Grasp/Reach Cortex

#### Electrode-Level Representation of Attempted Hand/Arm Movements

To examine the spatial distribution of movement tuning across cortical arrays, we implemented a cued movement task in which the study participant (RP2) attempted five distinct hand/arm movements in an instructed delay paradigm (Fig. 8A). Smoothed SBP activity from example electrodes revealed clear, distinguishable tuning to 5 relevant movement types (Fig. 8C). This analysis revealed significant decoding performance across a substantial fraction of electrodes on each array. Array yields, defined as the proportion of electrodes with better-than-chance decoding accuracy ranged from 54.8% (6V array) to 95.3% (AIP array) (Fig. 8D).

**Figure 7.**
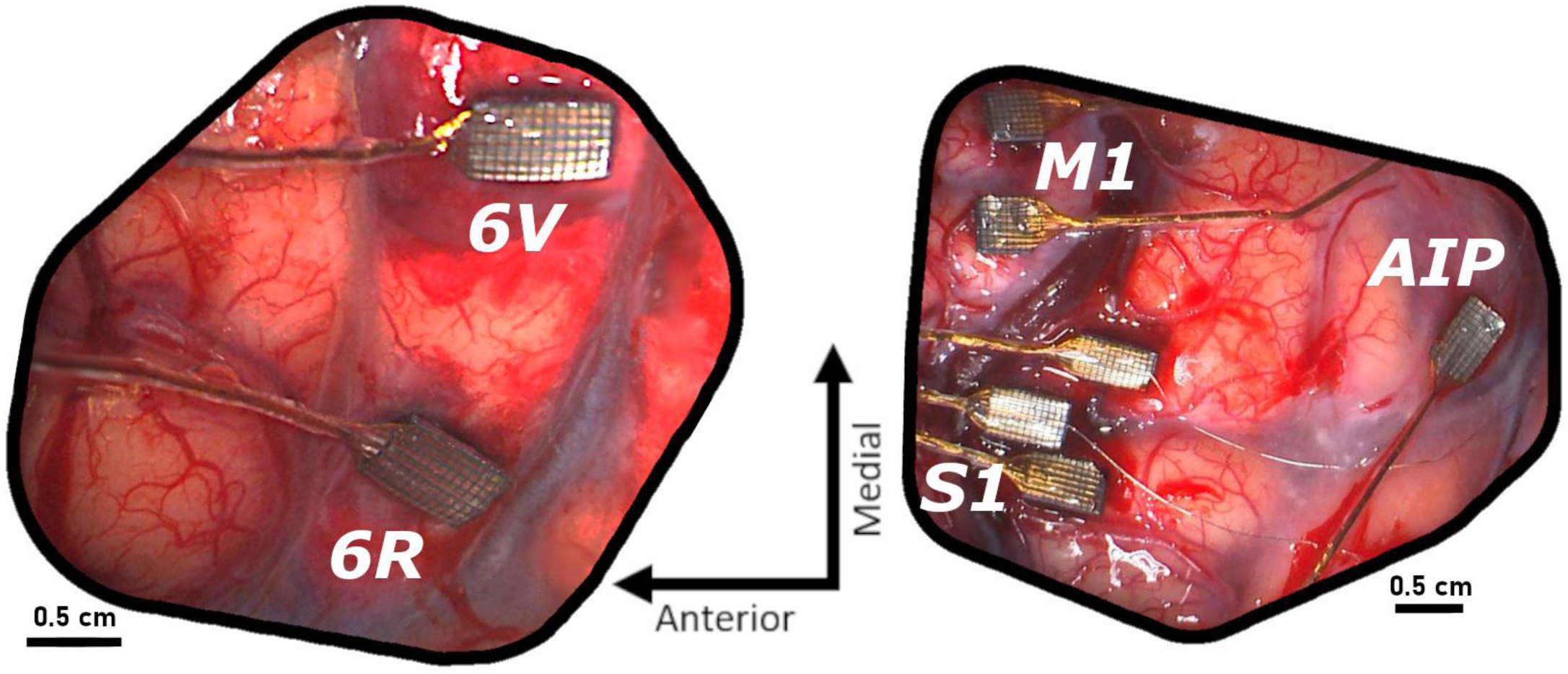
Intracortical implant sites. Left) Actual implant locations of the 6V and 6R arrays aligned with the view from Figure 6A,C. Right) Actual implant locations of the S1, M1, and AIP arrays aligned with the view in Figure 6B,D.

**Figure 8.**
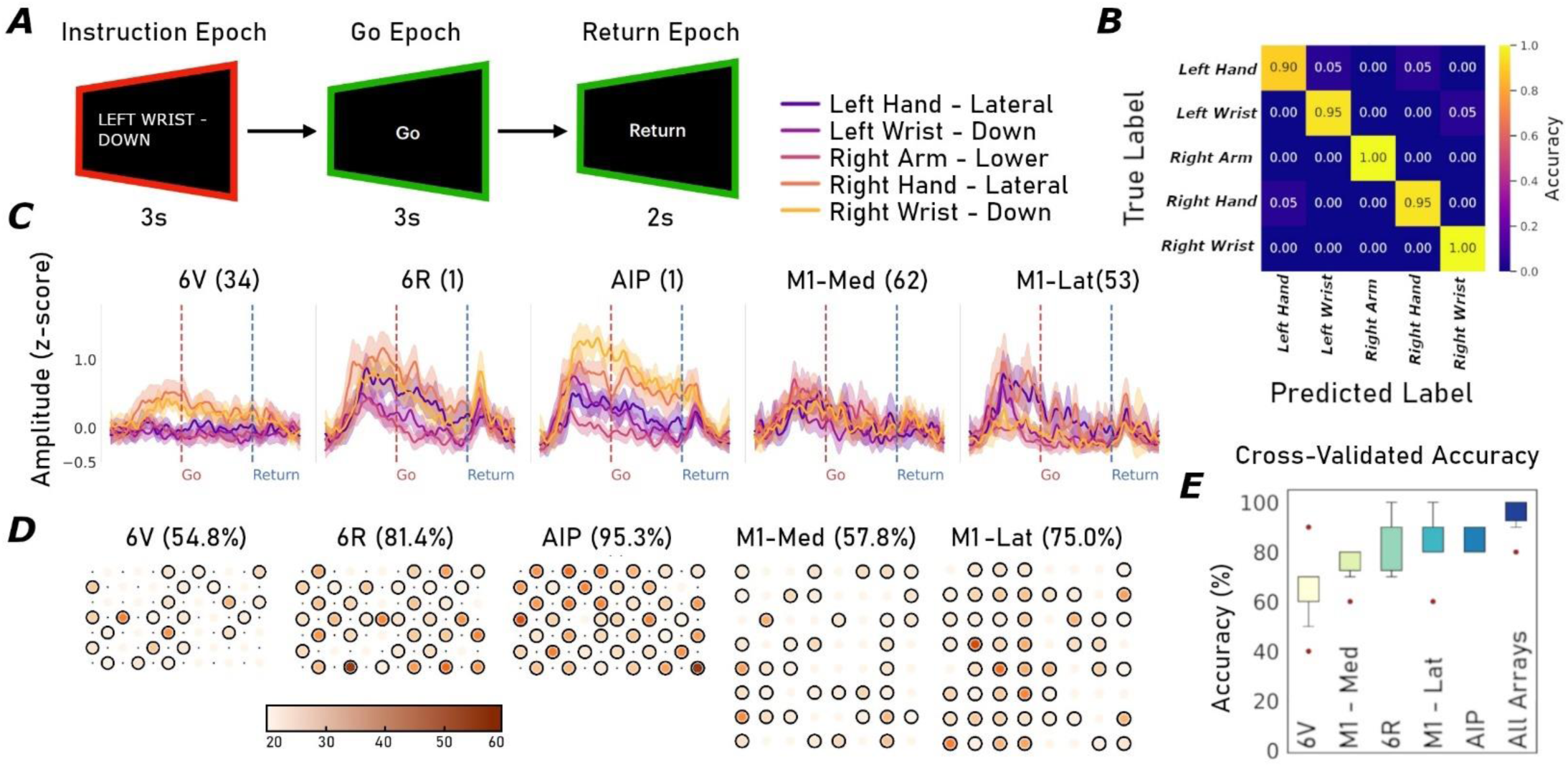
Neural tuning to bilateral hand/arm movements. **(A)** Cued movement task. Each trial consisted of three sequential epochs: an instruction cue indicating the movement to attempt, a go phase during which the participant executed the attempted movement, and a return phase in which the participant returned to a resting posture. **(B)** Confusion matrix showing decoding performance for the combined-array model across the five movement classes. **(C)** Single-electrode tuning. Example spike band power (SBP) features from individual electrodes (electrode number shown in parentheses) showing distinguishable time-varying responses to different movement types across cortical arrays. **(D)** Electrode-level decoding performance. Heatmaps showing the classification accuracy achieved using SBP features from individual electrodes. Color indicates decoding performance (darker is better), and array yield reflects the proportion of electrodes with accuracy significantly above chance. **(E)** Classification accuracies for each array individually, and for all arrays combined.

#### Complementary Representations Across Arrays

We next quantified array-level movement representation by first selecting optimal subsets of electrodes from each array using a greedy forward selection algorithm. Electrode subsets were combined across arrays to compute and visualize movement representation in a reduced dimensional space using principal component analysis. Next, we trained array-specific SVM classifiers using only the optimal electrode subsets. Consistent with the single-electrode results, all arrays demonstrated robust population-level encoding of movement types, with classification accuracies ranging from 65% (6V) to 86% (AIP) (Fig. 8E). This embedding revealed clear separation between movement conditions, indicating strong and structured encoding at the population level (Fig. 8B).

To assess whether different cortical regions encode movement types complementarily or redundantly, we combined the optimal electrodes from all arrays into a joint classifier. If cortical areas encode movements redundantly, no improvement in decoding accuracy would be expected. However, if regions contain distinct, complementary information, decoding accuracy should improve.

Indeed, we observed a substantial increase in classification performance—from a single-array best accuracy of 86% (AIP) to 96% when using combined electrodes—demonstrating that each region contributes some unique information about the cued movements (Fig. 8B,E and Supplementary Figure 3) These findings highlight the value of incorporating multi-regional neural signals for enhancing motor decoding in rehabilitation applications.

## Discussion

This study aimed to optimize intracortical microelectrode array placement for reach- and grasp-related motor decoding. We integrated functional imaging, anatomical mapping, awake mapping, and surgical feasibility to identify key implantation targets.

Using task-based fMRI and Quicktome-based anatomical mapping, we identified accessible locations in the anterior intraparietal area (AIP) and ventral premotor cortex (PMv) that showed grasp-related activity and connectivity. This approach improves upon previous methods that relied solely on anatomical landmarks. Implanting these locations, along with their connected primary motor and sensory cortices, will enable the decoding of more network-based reach- and grasp-related information. This information can then be used in brain-controlled functional electrical stimulation systems to restore arm and hand function in individuals with spinal cord injury.

### Distinguishing Grasp- and Reach-Related Activation

Although the broader goal of the ReHAB clinical trial is to restore both reach and grasp functions, our priority for electrode targeting was to identify grasp-related cortical areas. Therefore, we designed our fMRI protocol to separate grasping and reaching tasks, with the aim of distinguishing regions preferentially involved in grasp control.

However, the resulting activation patterns showed high consistency between grasp and reach conditions across all participants. While we observed areas with greater activation for grasp than reach in test subject 1 and participant RP2, this pattern was less apparent in test subject 2. The general similarity across conditions for all participants may stem from the interpretation of task instructions. Specifically, participants were not given explicit guidance on whether to imagine first reaching toward the object before grasping or to start from a pre-positioned hand and only imagine the grasp itself. This variability in mental imagery may have contributed to a lack of distinction between grasp- and reach-related activation.

Despite this, we still observed robust overall activation of reach- and grasp-related regions. Given that both functions are critical for the neural control of arm and hand movements, this provided us with viable and relevant implant regions regardless of the distinct separation between reach and grasp. Future investigations could refine the task paradigm by implementing more precise instructions or distinct movement sequences to enhance the differentiation between reach and grasp specific activation patterns.

### Comparison with Previous Implantation Studies

Our study’s methodology represents a direct evolution of the foundational approaches used to establish grasp-related areas as viable BMI targets. Previous studies from our lab and others have successfully guided implantations using anatomical landmarks,^37^ laying the essential groundwork for targeting high-level association cortices like AIP. A key insight gained from this body of work is the challenge posed by natural, inter-subject variability in functional anatomy. For example, our group previously observed unexpected auditory responses from an array targeted for AIP based on sulcal landmarks (unpublished data). ^37^

The multi-modal approach in our current study helps to explain and address this earlier finding. For participant RP2, our fMRI results showed that the classical anatomical landmark for AIP did indeed contain a subregion with strong auditory responses similar to our previous participant. Importantly, the fMRI also identified a distinct, adjacent subregion that was selectively active during imagined grasping without auditory confounds. By incorporating an auditory-contrast fMRI task, we were able to prospectively distinguish these two functional zones. This allowed us to refine the anatomical target and place the AIP array with greater functional precision, a step that would not have been possible with anatomical guidance alone. This example demonstrates how individualized functional mapping serves as a powerful complement to anatomical targeting, enhancing the potential for successful outcomes by ensuring implant locations are optimized for the intended BMI application.

### Implications and Future Directions

This study demonstrates a multi-modal approach to optimizing intracortical microelectrode array placement, integrating MRI-based anatomical mapping, fMRI-guided functional localization, and preoperative 3D surgical modeling. By incorporating task-based fMRI alongside Quicktome’s structural connectivity analysis, we improved implantation precision while reducing the risk of misplacing electrodes in non-motor areas. As brain-controlled upper limb functional electrical stimulation systems continue to evolve, integrating more diverse cortical targets beyond primary motor cortex will be crucial for advancing neuroprosthetic control of grasping and reaching in individuals with SCI. Future applications may extend this approach to other motor or sensory networks relevant for neuroprosthetic control.

## Data Availability

All data produced in the present study are available upon reasonable request to the authors

## Data availability

Data or materials that support the findings of this study are readily available from the corresponding author upon reasonable request.

